# Antimicrobial resistance and treatment failures in symptomatic and asymptomatic *Mycoplasma genitalium* infections in São Paulo, Brazil: Evidence from a cross-sectional study of sexual health clinic attendees

**DOI:** 10.1101/2025.04.14.25325535

**Authors:** Victor Cabelho Passarelli, Flora Goldemberg, Carolina Lázari, Gabriela Gaspar Carnevale, Mateus Vailant Thomazella, Sophia Chamarelli, Danilo Gennari, Caroline Pedrozo, Camila Melo Picone, Angela Carvalho Freitas, Gwenda Hughes, Silvia Figueiredo Costa

## Abstract

**OBJECTIVES:** We aimed to describe the epidemiology, antimicrobial resistance pattern, and treatment outcomes of symptomatic and asymptomatic *Mycoplasma genitalium* (MG) infections in patients from a public HIV/STI clinic in São Paulo, Brazil.

**METHODS:** This cross-sectional study ran from July 2023 to September 2024. Patients tested for bacterial STIs were invited to participate by consenting to sample storage for resistance analysis. Clinical management followed standard care at the clinician’s discretion, and data were retrospectively analyzed. MG positive samples were analyzed for macrolide and quinolone resistance associated mutations, respectively in the region V of the 23s rRNA and the ParC genes, using Allplex Azi-R and Moxi-R assays (Seegene, Inc., Seoul, Korea).

**RESULTS:** MG prevalence was 12.5% (47/376) in the study population and 66.0% (31/47) were asymptomatic infections. Mutations associated with macrolide (MDRM) and quinolone resistance were detected in 78.8% (26/33) and 21.1% (8/38) of the MG-positive samples, respectively. MDRM were significantly more common among symptomatic than asymptomatic patients (100.0% (13/13) vs 65.0% (13/20); p=0.016). Among untreated asymptomatic patients, 50.0% (3/6) cleared the infection without antibiotics. Treatment for 39.5% (15/38) included single-dose azithromycin, according to current national guidelines. Treatment failures were more common in the symptomatic group (91.7% (11/12) vs 40.0% (6/15); p=0.005).

**CONCLUSION:** Macrolide and quinolone resistance were common in *Mycoplasma genitalium* samples from an STI clinic in São Paulo, Brazil, with frequent treatment failures among symptomatic patients. Current Brazilian guidelines should be urgently revised. Screening and treating asymptomatic MG cases may promote antimicrobial overuse without clear benefits, as spontaneous clearance may be common. Larger studies are needed to guide management and clarify the clinical relevance of asymptomatic MG infections.

**KEY POINTS:** *What is already known on this topic:* *Mycoplasma genitalium* is common in vulnerable populations and poses a risk to antimicrobial resistance worldwide.

*What this study adds:* Detection of high rates of macrolide and fluoroquinolone resistance in *Mycoplasma genitalium* in Brazil, along with frequent spontaneous clearance in asymptomatic cases.

*How this study might affect research, practice or policy:* Findings support an update of Brazilian treatment guidelines and highlight the need for further investigation into the clinical relevance and management of asymptomatic infections.

## INTRODUCTION

*Mycoplasma genitalium* (MG) poses a significant challenge to global sexual health due to its high prevalence among vulnerable populations [1], as well as scarce treatment options and rising antimicrobial resistance (AMR), particularly to macrolides [1-4]. Surveillance is therefore of utmost importance.

In Brazil, however, surveillance for MG and AMR are absent. To date, only one study has reported an MG prevalence of 8.3% in men with urethral discharge across Brazilian regions [5], without further data on clinical or resistance profiles.

To address such gaps, we analyzed clinical and laboratory data from patients tested for STIs at our outpatient clinic in São Paulo, Brazil, evaluating the prevalence of macrolide and fluoroquinolone resistance in MG-positive samples, and treatment outcomes.

## METHODS

### Setting

Serviço de Extensão ao Paciente HIV/AIDS from the Hospital das Clínicas da Faculdade de Medicina da Universidade de São Paulo (“SEAP-HCFMUSP”) is a STI/HIV clinic in São Paulo, Brazil, which serves around 400 HIV-PrEP users and 3400 people living with HIV. All HIV-PrEP patients at our STI clinic are offered screening for bacterial STIs as part of their routine care. All other patients are tested only upon the presence of symptoms.

### Study design and study population

This cross-sectional study ran from July 2023 to September 2024. All patients tested for bacterial STIs were invited to participate by consenting to sample storage for resistance analysis. Clinical management followed standard care at the clinician’s discretion, and data were retrospectively analyzed.

### Sampling and laboratory procedures

Oropharyngeal, rectal and urine samples were self-collected by participants, placed in transport medium tubes and transported within 24 hours to our reference laboratory where they were initially tested for MG, *Neisseria gonorrhoeae (NG), Chlamydia trachomatis (CT) and Trichomonas vaginalis (TV)* with the Alinity m STI Assay (Abbott - USA), according to manufacturer’s instructions.

Positive samples for MG were subsequently analyzed for macrolide and quinolone resistance associated mutations, respectively in the region V of the 23s rRNA and the ParC genes, using the Allplex MG AziR Assay (Seegene, Inc., Seoul, Korea) and Allplex MG MoxiR Assay (Seegene, Inc., Seoul, Korea), according to manufacturer’s instructions. Data were analyzed using Seegene viewer software version 3 (Seegene, Inc., Seoul, Korea).

### Statistical analysis

Data were summarized using means, ranges, and percentages. Group comparisons used Student’s t-test for quantitative and Chi-squared or Fisher’s exact tests for categorical variables. Statistical significance was set at P < 0.05. Analyses were performed using Free Statistics Software (version 4.0).

## RESULTS

### Study population

Among 481 patients tested at the clinic during the study, 376 (78.2%) agreed to participate. Most of these (342/376; 91.0%) were recruited from our 400 registered HIV-PrEP users. Detailed demographics are shown in the table.

MG prevalence was 12.5% (47/376) in the study population. Coinfection with NG and CT occurred in 19.1% (9/47) and 4.3% (2/47) of the MG cases, respectively.

### Drug Resistance Mutations (DRM) in M. genitalium

Amplification and detection of DRM was possible for 70.2% (33/47) and 80.9% (38/47) for macrolides and fluoroquinolones, respectively. Unsuccessful amplifications were attributed to low bacterial load, degradation of frozen DNA during storage, and/or disparities in assay sensitivity between commercial platforms [1,10].

Of the samples analyzed, 78.8% had mutations associated with macrolide resistance (MDRM) and 21.1% had mutations associated with fluoroquinolone resistance (QDRM), table. We found MDRM to be significantly more common among symptomatic than asymptomatic patients (100.0% (13/13) vs 65.0% (13/20); p=0.016), but no significant difference in frequency of QDRM was observed between those groups (table). All samples presenting with QDRM also had MDRM (dual-resistance).

**Table.**
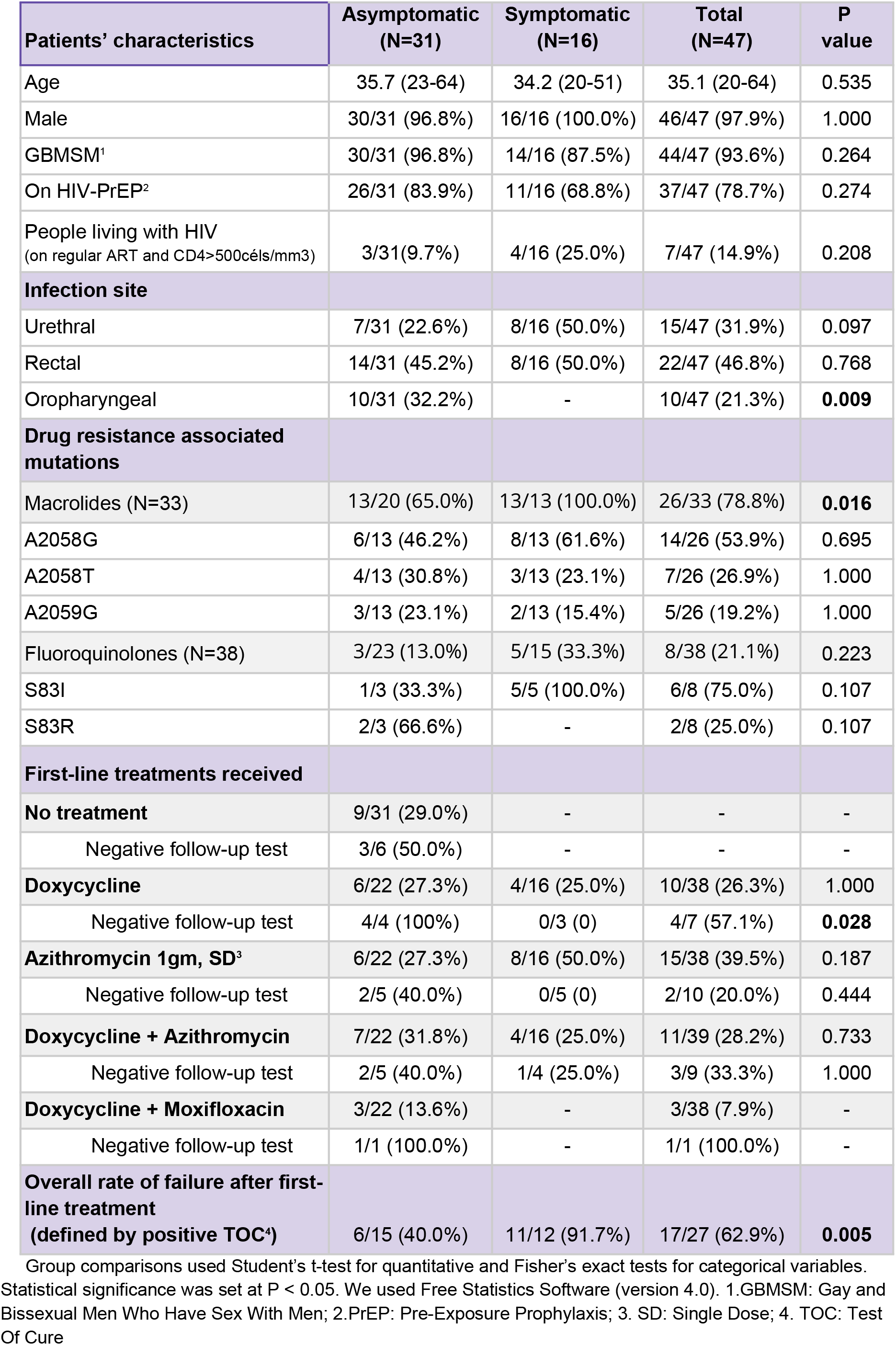
Characteristics of recruited patients diagnosed with MG, stratified by symptom status.

### Asymptomatic infections

Sixty-six percent (31/47) of positive tests were from asymptomatic individuals (table). Of these, 71.0% (22/31) received treatment and 29.0% (9/31) did not. None developed symptoms over the study period.

Among six untreated patients, 50% (3/6) cleared the infection without antibiotics; three others were lost to follow-up. First-line treatments were heterogeneous, with test of cure (TOC) available for 15 treated individuals (68.2%). Eradication rate varied (table), and six (40.0%) failed TOC. Only one, treated with doxycycline followed by moxifloxacin as second-line, cleared the infection. Those receiving doxycycline alone (2/4; 50.0%) or with azithromycin (1/4; 25.0%) did not clear the infection but remained asymptomatic, so no additional treatment was given. Two patients were lost to follow-up.

### Symptomatic infections

Thirty-four percent (16/47) of positive tests were from symptomatic patients. Half (8/16) presented with proctitis and half (8/16) with urethritis. None presented with oropharyngeal symptoms.

First-line treatments for symptomatic patients varied (table) and 91.7% (11/12) failed TOC, which was significantly higher when compared to asymptomatic patients (P=0.005), table.

Nine (81.8%) of those patients with a positive TOC received second-line treatments, including: doxycycline followed by moxifloxacin (4/9, 44.4%), azithromycin single-dose (3/9, 33.3%), doxycycline followed by azithromycin (1/9, 11.1%) and ciprofloxacin (1/9, 11.1%). All patients treated with doxycycline followed by moxifloxacin improved symptoms and had a negative TOC (4/4, 100%). While the other five patients also had symptom improvement, all failed TOC and had at least two persistent positive tests, each taken four months apart. However, they were not treated further as their symptoms had resolved. Two patients (18.2%) did not receive second line treatments, in spite of failing TOC, also due to symptom improvement.

## DISCUSSION

We found a high prevalence of *Mycoplasma genitalium* in our study population, similar to studies from different regions [1-4, 9-10] and highlighting the relevance of MG in urethral and rectal syndromes of sexual transmission in Brazil.

High rates of macrolide resistance-associated mutations were observed, consistent with global reports [1-3, 8-10]. In settings where resistance-guided therapy cannot be implemented, empirical azithromycin monotherapy may be inadequate, especially for vulnerable populations such as GBMSM [1]. Despite growing evidence of treatment failure and resistance emergence [2], Brazilian STI guidelines still recommend single-dose azithromycin as first-line treatment for uncomplicated MG infections [6]. Among symptomatic patients in our study—most of whom received azithromycin-based regimens— treatment failures were common, supporting the urgent need to revise national guidelines. Notably, resistance mutations were significantly more prevalent in symptomatic patients, arguably reflecting prior empirical macrolide use or higher bacterial loads [1], although this warrants further evaluation in larger studies.

Quinolone resistance associated mutations were also high, correspondingly with other clinical studies globally [2]. All patients who had MG infections with ParC gene mutations also had 23S gene mutations (dual-resistance), concordant with recent data from the UK and China [1,9].

In our study, most patients with MG (66.0%) were asymptomatic and received varied antibiotic treatments due to lack of specific guidelines—despite remaining symptom-free. Interestingly, a higher proportion of asymptomatic patients had a negative TOC after treatment compared to symptomatic ones. However, half of the untreated asymptomatic patients also cleared the infection, suggesting spontaneous resolution may be common. A recent study [8] showed azithromycin cleared MG infection in 68% of 38 cases, predominantly asymptomatic, in a scenario with high macrolide resistance prevalence - raising questions about the true impact of antibiotics in such a setting.

The clinical relevance of asymptomatic MG remains uncertain [1,3], and routine screening in high-prevalence settings may lead to unnecessary, inconsistent treatment and contribute to antimicrobial resistance [2,7]. Despite guidelines advising against screening asymptomatic individuals for MG [3,4], our clinic routinely does it because the only available molecular assay is a multiplex panel that includes it.

Brazil is a large middle-income country with limited access to molecular STI diagnostics, especially in public healthcare. Our clinic is the only Brazilian public STI center routinely testing for MG, so broader access to diagnostics and AMR surveillance tools are essential. Moreover, moxifloxacin is expensive and unavailable in the public health system, highlighting another obstacle to effective treatment in Brazil. Alternatives like pristinamycin [3] currently lack regulatory approval and are therefore unavailable.

Our research has limitations. The study population represented a small proportion of patients from one HIV/STI outpatient clinic in São Paulo, leading to selection, availability and sampling biases and limiting the strength and generalisability of our findings. These should be confirmed with larger samples from different regions. Also, unsuccessful amplification with some samples might have overestimated or underestimated the prevalence of DRM.

Our work adds value to current scientific knowledge about epidemiology, clinical characteristics and antimicrobial resistance profile of MG in Brazil. As the first study to explore this in detail, it collaborates to fill the gaps in surveillance and management in the country.

## CONCLUSION

Macrolide and quinolone resistance were common in *Mycoplasma genitalium* samples from an STI clinic in São Paulo, Brazil, with frequent treatment failures among symptomatic patients. Current Brazilian guidelines should be urgently revised. Screening and treating asymptomatic MG cases may promote antimicrobial overuse without clear benefits, as spontaneous clearance may be common. Larger studies are needed to guide management and clarify the clinical relevance of asymptomatic infections.

## Data Availability

All data produced in the present study are available upon reasonable request to the authors

## STATEMENTS

### Author contributorship

SFC conceived the study and is the guarantor of the work.

VCP, GH, ACF and SFC contributed to study conception, study design and interpretation of data.

VCP, CMP, ACF and FG contributed with data collection.

CL, GGC, MVT, SC, CP and DG performed laboratory analysis and resistance profiling. VCP, GH and SFC drafted the initial manuscript.

All authors critically revised the manuscript for intellectual content and approved the final version for submission.

### Ethics approval

This study was approved by the Local Ethics Committee (CAAE: 77048124.2.0000.0068).

### Patient consent

All patients who agreed to participate signed a written informed consent form.

### Data sharing

Data available upon reasonable request.

## Acknowledgments

This work was accepted for oral presentation at the ESCMID Global 2025 conference, to be held in Vienna, Austria 12-15th April, 2025.

Authors would like to thank the entire team at SEAP-HCFMUSP for assistance.

